# Retrospective study of the first wave of COVID-19 in Spain: analysis of counterfactual scenarios

**DOI:** 10.1101/2021.02.16.21251832

**Authors:** Benjamin Steinegger, Clara Granell, Giacomo Rapisardi, Sergio Gómez, Joan T. Matamalas, David Soriano-Paños, Jesús Gómez-Gardeñes, Alex Arenas

## Abstract

One of the most important questions on the COVID-19 pandemic is ascertaining the correct timing to introduce non-pharmaceutical interventions (NPIs), based mainly on mobility restrictions, to control the rising of the daily incidence in a specific territory. Here, we make a retrospective analysis of the first wave of the epidemic in Spain and provide a set of useful insights to optimize actions in the near future. We have reconstructed the exposure times, from infection to detectability, to correctly estimate the reproduction number R_t_. This enables us to analyze counterfactual scenarios to understand the impact of earlier or later responses, decoupling containment measures from natural immunity. Our results quantify the differences in the number of fatalities for earlier and later responses to the epidemic in Spain.

**Teaser:** *“We propose a backward analysis of pandemic incidence in a region to determine the correct timing of authorities’ non-pharmaceutical interventions to fight COVID-19”*

---

Spain was among the strongest hit countries globally during the first wave with officially 28.000 fatalities caused by COVID-19, in excess deaths even 40% more, and an attack rate between 4% and 5% (1). Surprisingly, the Spanish authorities implemented very stringent containment measures, which contrasted with the high fatality and attack rates. The national government implemented the first lockdown on March 15, which was reinforced on March 29 until April 12. The confinement was then gradually lifted from May 2 onward.

In short, the non-pharmaceutical interventions NPIs issued by the authorities, all together with physical distancing, face masks, and hands hygiene adopted by the population, were sufficient to mitigate the daily infections. Therefore, a natural retrospective question arises: how to identify the impact of the various NPIs implemented at the timing they were implemented. The answer is essential to design the future mitigation strategies of COVID-19, as it has been done for China or Italy (2,3).

To assess the impact of NPIs, one can either directly analyze the epidemiological data such as case numbers, hospitalizations, and fatalities (4) or perform model-based inference (5, 6) to extract cornerstone conclusions. Here, we blend both approaches. Analyzing the epidemiological data allows us to propose a hypothesized functional form for R_t_ during Spain’s first wave. Then, we combine this information with a model-based approach to infer the behavior of R_t_. With this analysis, we can explore counterfactual scenarios.

## Results

One of the most pressing difficulties when analyzing an epidemic’s impact retrospectively is estimating the unobserved past incidence of the infection (7). Estimating the past incidence of COVID-19 relies on the accurate determination of the period’s distribution between the exposure to the virus to affliction (or detectable symptomatic infection).

In Spain, the ISCIII provides a time series with symptom onset dates for reported cases (8). Using current estimates of the incubation period, we can infer the daily infections using a non-parametric back-projection method based on a maximum likelihood estimation (7) (see Materials and methods).

In Fig. 1A we show the results of the estimated infections for the Comunidades Autónomas (CCAA) in Spain, as well as for the whole country, together with the corresponding official data for symptoms onset and reporting date. The substantial delays between infection and report are evident for all regions, as corroborated in Fig. 1B showing the day when the infections and reported cases peaked, andthe difference between them. On average, infections and reported cases peaked with a 15 days difference (sd: 3). We find the maximal difference of 20 days for Catalonia, together with Castilla La-Mancha.

**Figure 1:**
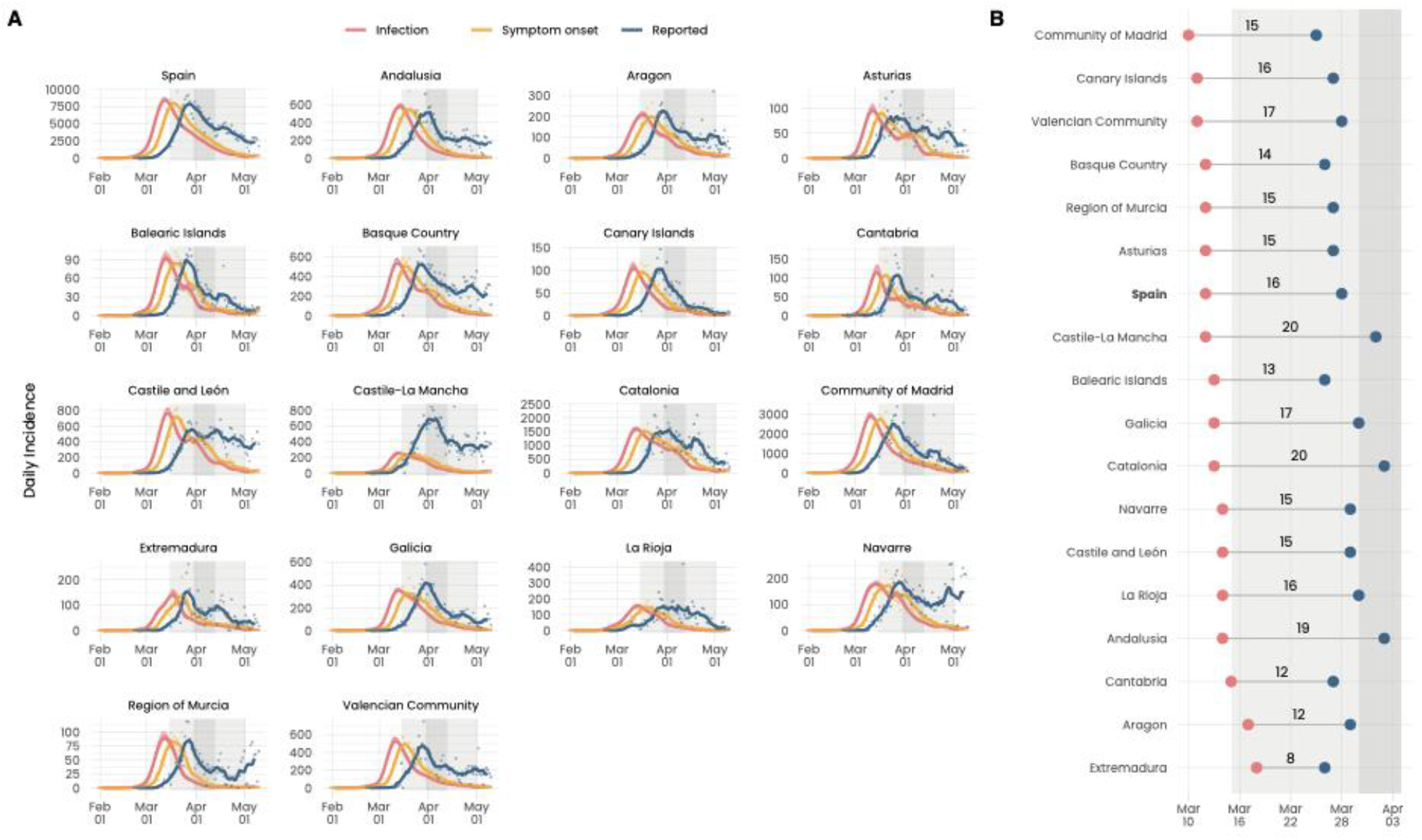
Reconstructed infections vs. reported notifications in Spain. (**A**) Time series for reported cases in blue and symptom onset in yellow for Spain and the CCAA. Solid lines indicate a centered, 7 day rolling average. In red we show the reconstructed exposure times. The shaded area in light gray indicates the lockdowns 1 and 3. In dark gray, we indicate the lockdown 2 where, in addition, all non essential economic activity was shut down. (**B**) Date on which the reconstructed exposure (red) times and reported cases (blue) peaked. Additionally, we explicitly indicate the delay between the two peaks. Shaded areas and bars around lines indicate 95% credible intervals in the entire manuscript.

Then, we estimated R_t_’s evolution in Fig. 2A using EpiEstim (9). In all CCAA, we observe a substantial reduction of R_t_ before the first confinement. Close to this first confinement, R_t_ is for the first time below one in almost all CCAA, and stayed mainly below one until the end of the confinement. Fig. 2B shows the day on which R_t_ was first below one in the CCAA. From March 13 to 21, R_t_ became smaller than 1 in all CCAA. On average, R_t_ crosses 1 on March 16 with a standard deviation of 2,1 days, which is consistent with other estimations (14, 15). The Community of Madrid, was the first CCAA to push R_t_ below 1 as a consequence of the implemented containment measures on March 9, before any other CCAA. Similar results are found for other CCAA (see SM for detailed information about the evolution across CCAA).

**Figure 2:**
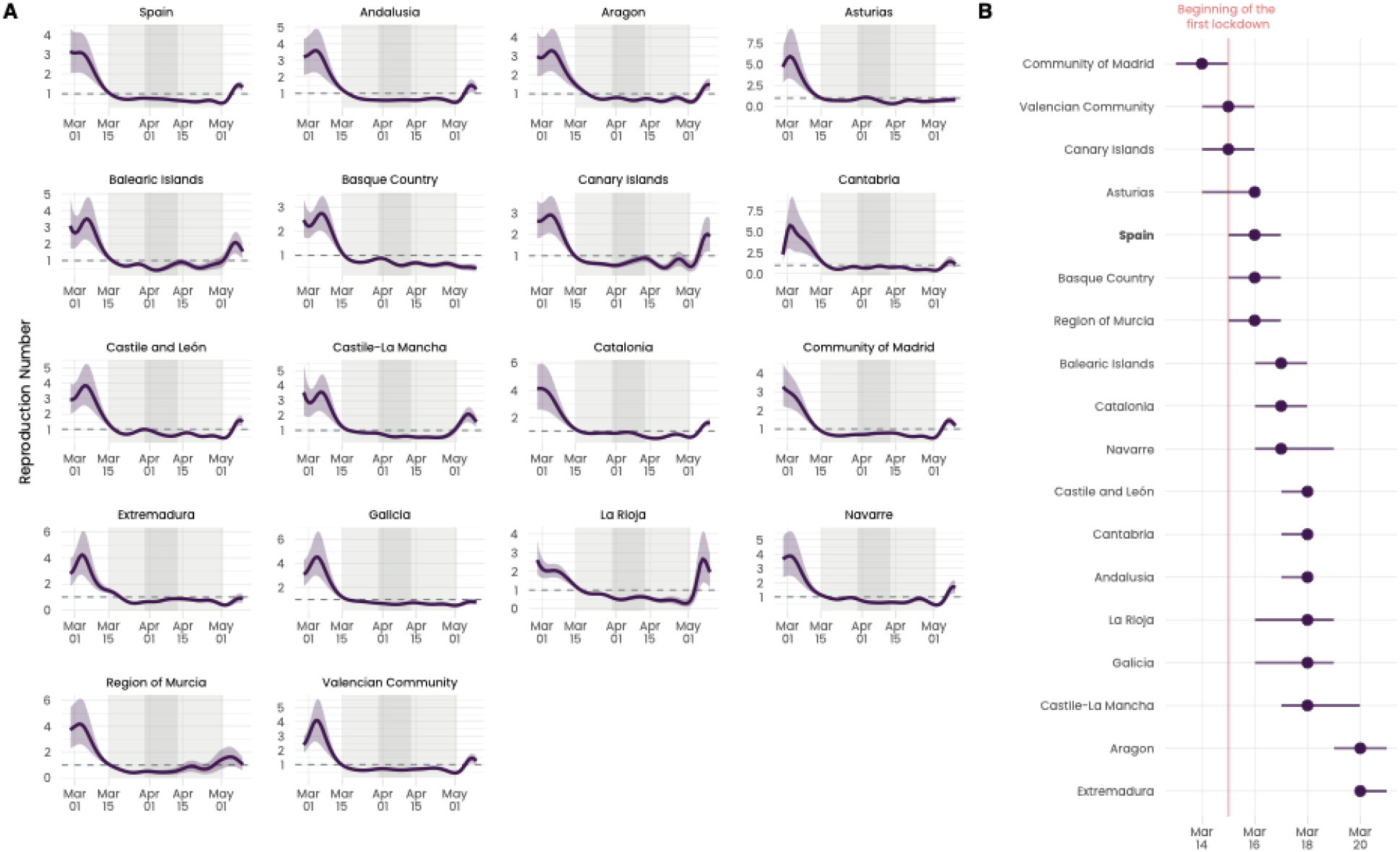
Estimation of R_t_ from reconstructed infections. (**A**) Estimation of R_t_for Spain and the CCAA from the reconstructed exposure times. The shaded area in light gray indicates the lockdowns 1 and 3. In dark gray, we indicate the lockdown 2 when all non-essential economic activity was shut down. (**B**) Chronological order for the date when R_t_ dropped first below 1 in Spain and the CCAA.

After obtaining the retrospective values of R_t_, and given that the NPIs impact starts with the behavior of people, we wonder when the epidemic response was initiated. The epidemic response should result in a breakpoint (17) in R_t_ after which we observe a substantial reduction of R_t_. To determine this breakpoint, R1, we separate (see Materials and Methods) R_t_’s evolution into three linear segments: the “free” spreading phase before restrictions, the initiation of the epidemic response, and the lockdown. The result is shown in Fig. 3A. The second breakpoint pretty much coincides with the implementation of the first lockdown by the authorities in Spain. More interestingly, the reproduction number starts already to decrease between March 5 and 6 (first breakpoint). The early decrease precedes the introduction of any containment measures, also on a regional level, as well as the mobility reduction, which started between March 9 and 10 (see Fig. S1).

**Figure 3:**
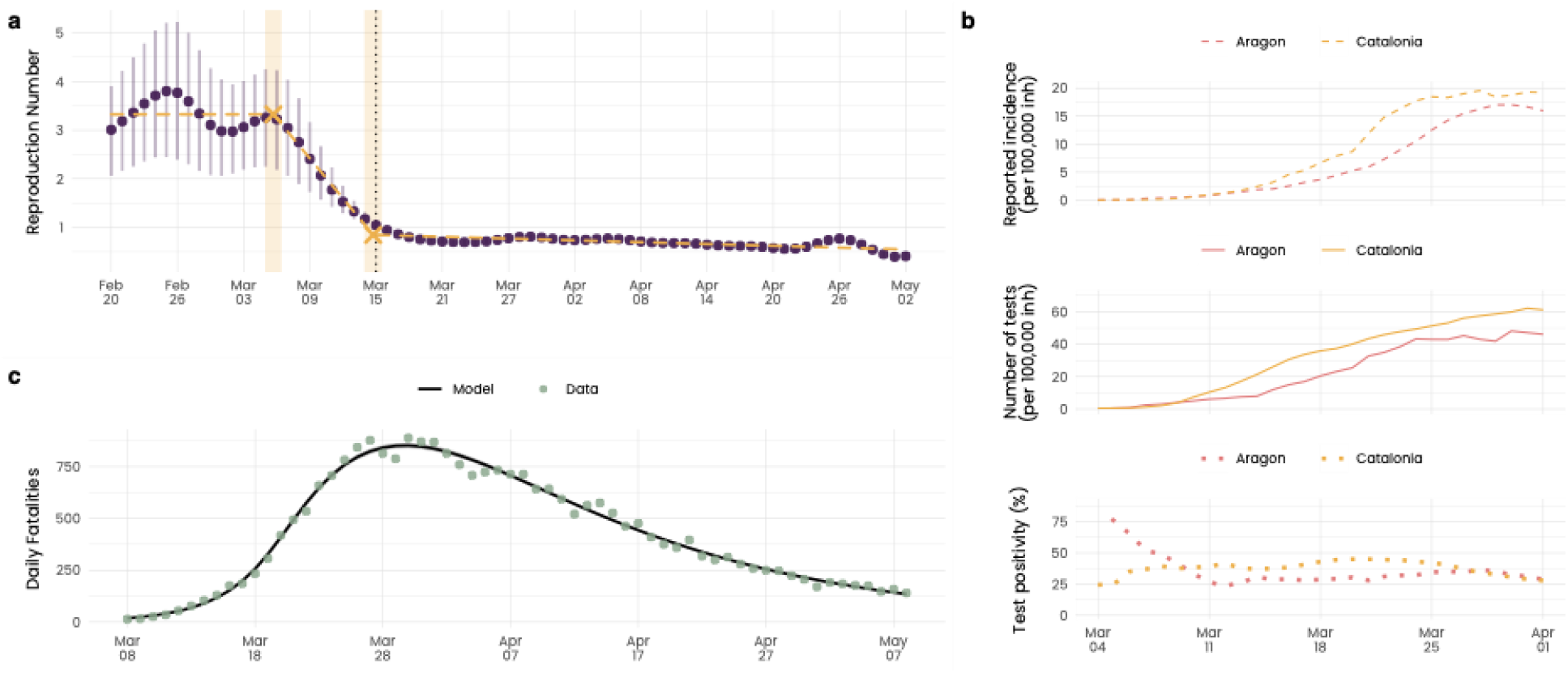
Early decrease of R_t_ and saturation of testing capacity. (**A**) Estimated evolution of R_t_in Spain. Yellow dashed lines and crosses indicate the identified segments and breakpoints, respectively, in the time series. We explicitly fixed three segments to separate the “free” spreading phase, the epidemic response, and the lockdown. (**B**) Time evolution of the per capita incidence (top), number of tests (middle) and test positivity (bottom) in Aragón (18) and Catalunya (19). While cases rise exponentially, the number of tests only increases linearly. Since test positivity does not significantly change, this strongly indicates saturation in test capacity. (**C**) Fit between the model and the daily fatalities.

However, this early decrease is found based on the reported case data. Therefore, the estimation of R_t_ is subjected to changes in the testing capacity and reporting criteria. The preceding decrease of R_t_ concerning mobility and containment measures could be due to a saturation of the test capacity, as illustrated in Fig. 3B for the CCAA Catalonia and Aragon. To corroborate this hypothesis, we compare the estimation with the most reliable epidemiological data in Spain, daily fatalities. In particular, we use our estimation to inform a simple, but robust, model (Methods Materials) about R_t_’s general form, and adjust it to the daily fatalities. The first breakpoint, BP, which indicates the start of the epidemic response, as well as the initial number of infected individuals, I0, are left as free parameters. To reduce the number of free parameters, we fix the second breakpoint on March 15, one of two break points in the time series for R_t_, Fig. 3A. Additionally, we assume a constant value R2 of R_t_ during the lockdown.

Figure 3C shows the fit of the model to the time series of daily fatalities. The parameters were calibrated as: I0 = 159(CI: 80-276), R1 = 4.75(CI: 4.3-5.3), R2 = 0.713(CI: 0.704 – 0.721), BP = 2020-03-12(CI: 2020-03-10-2020-03-13). Figure S2 presents the summary statistics. The corresponding evolution of R_t_ is shown in Fig.4A. The estimations of R1 and R2 lead to a doubling time and half-life time of 1.84(CI: 1.69– 1.99) days and 11.2(CI: 10.8– 11.6) days, respectively (27).

**Figure 4:**
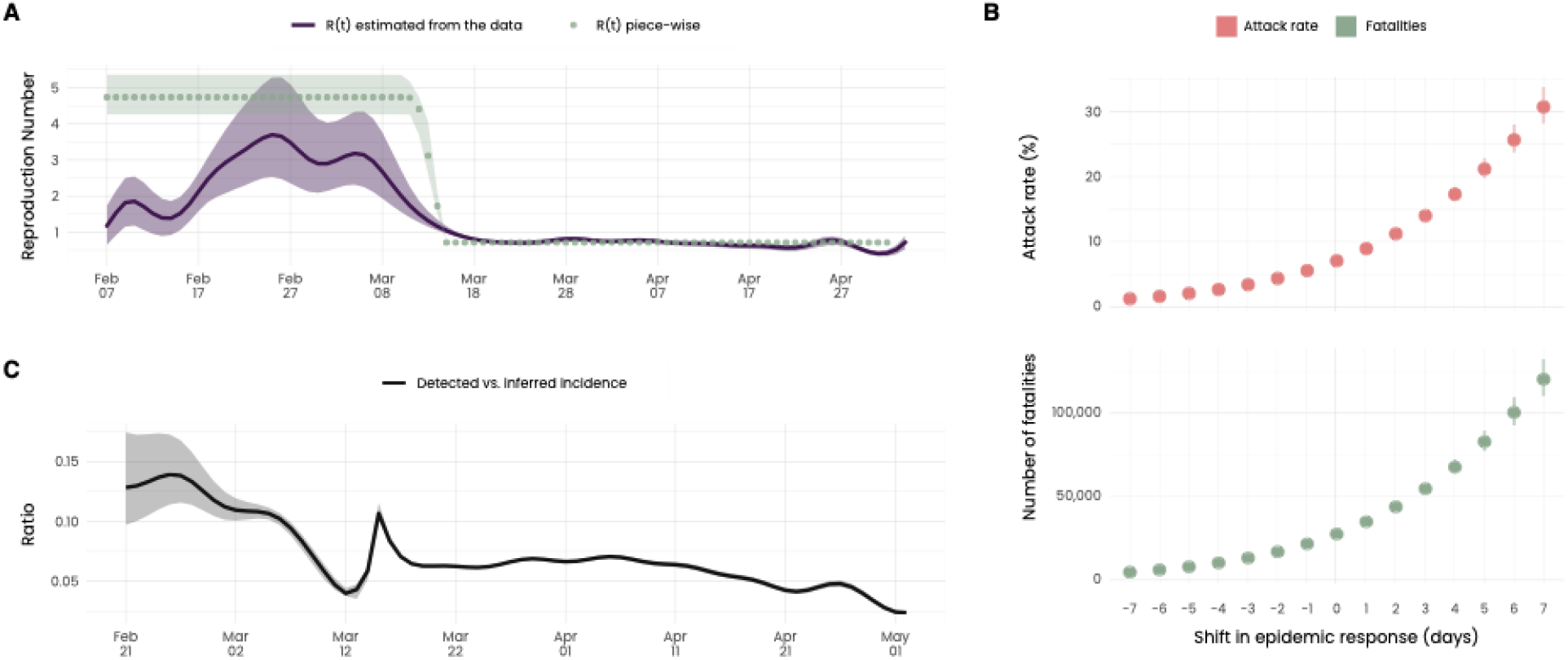
Underdetection and counterfactual scenarios. (**A**) Estimated R_t_ from the reconstructed exposure times together with the piecewise linear R_t_, which we inferred through a fit to the daily fatalities. (**B**) Attack rate (top) and total number of fatalities (bottom) as a function of the shift in number of days of the epidemic response. (**C**) Estimated ascertainment bias according to the ratio between the daily infections from the model and the reconstructed exposures from the reported cases.

In Fig. 4A, we compare the R_t_ estimated from official incidence data, with the one fitted to reported daily fatalities. The first observation is that the actual R_t_ seems to have been substantially higher than what can be estimated from the reported incidence data. Additionally, we observe a reduction in R_t_ between March 10 and 13 substantially later than inferred from the reported data. Importantly, this indicates a decrease in R_t_, preceding the implementation of the national lockdown. This seems to be the fingerprint of a spillover effect (16) from the earlier introduction of local restrictions as in Madrid, for example. These earlier restrictions raised the individual awareness of the population as indicated by an increase of COVID-19 related searches in Google from March 9 onward (see Fig. S3).

Figure 4C shows the estimated ascertainment bias according to the ratio between the daily infections from the model and the exposure times. Our results indicate that, initially, ascertainment was only around 10%. Additionally, ascertainment continuously deteriorated towards the first confinement, when it almost reached 5%. This decreasing ascertainment indicates that the ramping up of the test capacity could not keep up with the propagation of the epidemic.

Finally, the inferred form of R_t_ serves us to build counterfactual scenarios. We evaluate the accumulated fatalities, as well as the attack rate, if the epidemic response (including confinement, an earlier adoption of social distancing and the specific measures taken by the CCAA) had occurred earlier or later by shifting the function of R_t_.

Figure 4B presents the total attack rate and accumulated fatalities if the entire epidemic response had occurred some days earlier/later. For a 7 days earlier response, we find an attack rate of 1.2% (CI: 1.1– 1.4%) and a total number of fatalities of 4,782 (CI: 4,146-5,503). In contrast, supposing the epidemic response 7 days later, we find an attack rate of 30.8% (CI: 28.2-34.0%) and a total number of fatalities of 120,353 (CI: 110,165-132,545). Due to the high R_t_ in Spain, which translates into a minimal doubling time, any delay in the epidemic response would have had drastic consequences.

## Discussion

Our results indicate the existence of a substantial delay between exposure and notification in Spain. An average delay of 15 days proves how late individuals with symptoms were tested, the substantial test turnaround time, and the significant delay to be notified. Such pronounced delay hinders an actual evaluation of the epidemiological situation and therefore impedes effective management of the epidemic response. For illustration, as the first confinement was implemented, our results indicate that the daily infections had already peaked. Similarly, on March 28, all non-essential economic activity was shut down despite still rising case numbers. However, we find that daily infections had already peaked more than 10 days before in most CCAA.

Besides the homogeneous, impactful containment measures implemented on a national level, we found that the earlier introduction of containment measures on a regional level led to differences when the daily case numbers first stopped growing for upto a week. In general, the CCAA,which first pushed R_t_ below 1,coincides with the ones that implemented containment measures the earliest. The most prominent example is the Community of Madrid. Despite the regional heterogeneity, the first confinement on March 15 fell into the range when R_t_ became smaller than 1.

By fitting a hypothesized functional form of R_t_ to reproduce the evolution of daily fatalities, we found that R_t_ started decreasing between March 10 and 13. The first containment measures taken from March 9 onwards and the corresponding mobility reduction which started around March 10, fall into this range. This reduction of R_t_, before the first confinement, highlights the role of individual awareness of the population, a factor whose consideration is crucial (16). Nonetheless, the observation based on reconstructed infections contrasts with the reported data, which indicates a constant decrease in R_t_ from March 5 onwards. Available data from Aragón and Catalunya indicates that this earlier decrease can be attributed to saturation in the testing capacity. Similarly, the higher R_t_ value inferred from the model suggests that the expansion of test capacity could not keep up with the propagation speed of the epidemic in Spain. Accordingly, constantly fewer cases were detected towards lockdown before eventually only capturing approximately 5% of cases. In contrast, Althaus et al. find the opposite for Switzerland, where an increasing percentage of cases was detected towards lockdown (15). A decreasing detection rate, a substantial delay between exposure and notification, together with a doubling time of about 1.8 days, hampered the evaluation of Spain’s epidemiological situation and may have led to an underestimation regarding the severity of the situation.

Our counterfactual scenarios indicate that only a minimal delay in the epidemic response leads to a substantially different outcome. For an epidemic response 7 days earlier, we find approximately 5,000 fatalities and an attack rate of 1%, while a 7 days later response results in 120,000 fatalities and an attack rate of 30%. In this sense, the epidemic response and NPIs prevented many fatalities. Nevertheless, an earlier introduction of these measures would have substantially reduced COVID-19 related mortality.

## Data Availability

Only public data was used.

## Acknowledgments

The authors acknowledge Prof. Dan Larremore and Prof. Miguel Hernán for their helpful comments on the manuscript.

## Author contributions

A.A., S.G. and J.G.-G. are joint senior authors. B.S. and A.A. designed the project. B.S., J.T.M., G.R. and D.S.-P. collected data. B.S., J.T.M., D.S.-P., G.R. and C.G. analyzed the data. B.S., J.T.M., D.S.-P., G.R., S.G, C.G. J.G.-G. and A.A. interpreted the results. B.S., J.T.M., D.S.-P., G.R., S.G, C.G. J.G.-G. and A.A. wrote the manuscript.

## Data and materials availability

All data is available in the main text or the supplementary materials.

## Supplementary Material for

### Materials and Methods

#### Reconstruction of exposure times

To reconstruct the exposure times in the different CCAA, we use the function backprojNP (*17*) from the surveillance package (*18, 19*) in R. The method, initially proposed by Becker et al. (*7*), infers the expected number of exposures given the probability mass function of the incubation period through a maximum likelihood approach.

Credible intervals are calculated based on a bootstrap procedure (*20*). We fixed the smoothing factor *k* = 6, which corresponds to a centered rolling average of 7 days. The bootstrap procedure made use of 1000 samples (*B* = 1000). The incubation period distribution was fixed as a gamma distribution with mean 5.2 days and standard deviation 2.8 days (*21*). The time series with the symptom onset is provided by the Centro Nacional de Epidemilogía (*22*).

#### Estimation of *R*_*t*_

From the median incidence, obtained by the reconstruction of the exposure times, we estimate the evolution of *R*_*t*_ using the EpiEstim package (*9, 23*). Our interest lies in the impact of the NPI. Accordingly, we estimate the instantaneous *R*_*t*_ (*24*) rather than the case *R*_*t*_ (*25*), since the former reflects changes in the transmission dynamics more abruptly.

For the infectivity function, we choose the generation time estimated by Ganyani et al. (*12*). To be more precise, we assume a generation time following a gamma distribution with mean 5.2 (CI : 3.78 – 6.78) days and standard deviation 1.72 (CI : 0.91 −3.93) days. This generation time distribution corresponds to the estimation by Ganynai et al. for Singapore while assuming the same incubation period distribution (*21*) as we do in the reconstruction of the exposure times. We assume a standard deviation for the mean and the standard deviation of the generation time of 1.0 days and 1.2 days, respectively. However, we bound the values for the mean and standard deviation by the estimations of Ganyani et al..

We fixed a centered rolling average of 7 days. To bootstrap the credible intervals, we took 100 samples of the generation time distribution and considered 100 posteriors for each of these samples (n1 = 100, n2 = 100).

#### Identifying segments of *R*_*t*_

To identify the linear segments we use the R package segmented (*13,26*). The method proposed by Muggeo implements a maximum likelihood approach using linear predictors. The credible intervals are obtained through bootstrap. As previously pointed out, we assume three segments of *R*_*t*_. An initial constant value R1 as the disease was spreading freely in Spain as well as two linear evolving parts corresponding to the decrease of *R*_*t*_ towards lockdown and the constant decrease observed during the lockdown.

#### Adjusting the model to the daily fatalities

To evaluate the validity of our estimation of *R*_*t*_, we fit a piece-wise linear *R*_*t*_ to the epidemiological data in Spain. We then compare this fit with our estimation of *R*_*t*_ from the exposure times. Unfortunately, the only reliable time series available are the daily fatalities (*27*). Both, hospitalizations and ICU occupation, were subject to various changes of reporting criteria from March until May 2020. As an example, CCAA switched between reporting the accumulated number of admissions and the current number of ICUs occupied without announcement. For this reasons, we refrain from using hospitalization data and focus on the daily fatalities.

We assume *R*_*t*_ to have three segments as in the previous analysis. Motivated by the identified segments and to reduce the number of parameters to fit, we fix the second breakpoint on March 15, the day the lockdown was implemented. It seems reasonable that, as the first lockdown was implemented, *R*_*t*_ reached a relatively stable value. In contrast, the first breakpoint BP, which initiates the decrease of *R*_*t*_ and thus the epidemic response, we leave as a free parameter. In this way, we are able to evaluate to which extent the decrease of *R*_*t*_ before lockdown actually took place. Additionally, we assume a constant value R1 and R2 of *R*_*t*_ before and after the second breakpoint.

We then inform an epidemic model with the assumed form of *R*_*t*_. Given *R*_*t*_, the generation time distribution *w*(*t*) and the size of the population *N*, the daily incidence *I*_*t*_ on day *t* evolves as (*9, 24*):

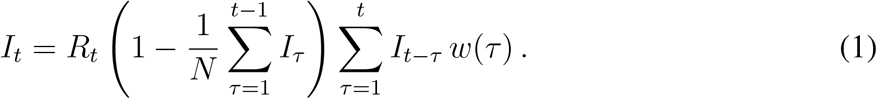

In the above equation, we decoupled the effect of containment measures and a reduction of *R*_*t*_ due to natural immunity. This separation serves us later to build the counterfactual scenarios. We use the same generation time distribution as for estimating *R*_*t*_ (*12*). From the daily incidence, we propagate the symptom onset as well as the daily fatalities through convolution (*28*). Given the incubation time distribution *P* (*t*) as well as the time from symptom onset to decease *D*(*t*), the daily number of individuals developing symptoms *S*_*t*_ and the daily fatalities *F*_*t*_ evolve as:

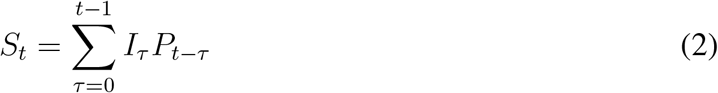

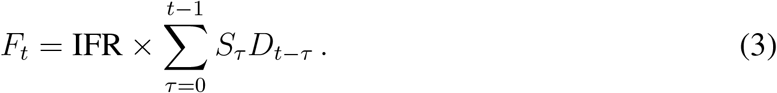

We fix the incubation time period distribution as for the estimation of the exposure times (*21*). We set the IFR as 0.83% according to the nationwide seroprevalence study (*29*). The average time from symptom onset to decease is fixed as 11 days according to the information by the public health authorities (*30*). Since no information is available on the type of distribution which fits best the data from symptom onset to decease, we make use of a gamma distribution (*5, 31*). We fix the shape factor (0.63) to match the interquartile range of the empirical distribution of 7 and 17 (*30*). In the Supplementary Text we perform a sensitivity analysis by considering a log-normal distribution.

Finally, in order to fit the model to the data, we make use of a MCMC approach. To be more precise, we use the DREAMzs sampler (*32, 33*) from the BayesianTools package (*34*). We use all flat priors, namely I0 ∈ [1, 1000], R1 ∈ [3, 8], R2 ∈ [0, 1] and BP between March 1st and 15. The number of iterations is fixed to 3 ·10^6^ with three chains. We fix the snooker update probability as 0.5. We use 20% of the iterations for adaption and discard the first 30% of the iterations. The posteriors and their correlation are shown in Fig. S2, whereas the actual fit is presented in Fig. 3C.

#### Counterfactual scenarios

To analyze what would have happened if the epidemic response had occurred earlier or later, we make use of the previous fit of the piece-wise linear *R*_*t*_. By shifting the breaking points, we can analyze straightforwardly the impact of an earlier or later response since we decoupled the containment from the effect of natural immunity. This approach is strongly inspired by Althaus et al. (*11*). We make all the simulations sampling 5000 parameter sets from the calibration’s posterior to bootstrap the credible intervals.

## Supplementary Text

### Heterogeneity in the epidemic response between CCAA

In the main text, Fig. 2B shows the precise day on which *R*_*t*_ was first below one in the CCAA. Here, we describe in more detail how this day correlates with the measures implemented in the CCAA. The Community of Madrid was the first CCAA to push *R*_*t*_ number below one. Madrid was the strongest hit CCAA early on and implemented first containment measures on March 9 before all other CCAA. Similarly, other CCAA, which first pushed *R*_*t*_ below one, were among the first to introduce containment measures such as Asturias (*35*), Valencian Community (*36*) or Basque Country (*37*). Interestingly, the last CCAA to push *R*_*t*_ below one, are in geographical proximity to Madrid such as Aragon, Castile-La Mancha, Castile and León, and Extremadura. This is consistent with a study by Mazzoli et al. that indicates how the local incidence strongly correlated with mobility in the early phase from and to Madrid, due to multi-seeding (*38*). Furthermore, Castile-La Mancha, Castile and León, and Extremadura have the highest percentages in Spain of their students attending university in the Community of Madrid; 27%, 14%, and 12%, respectively (*39*). Even though it is speculative, it seems reasonable that, after the closure of the universities, the outflux of students from Madrid to their hometowns added various seeds with mostly mild symptoms, accelerating the evolution of the epidemic in these regions.

### Cross-correlation between mobility and the regional *R*_*t*_

The SARS-CoV-2 epidemic led to a substantial reduction in mobility. Figure S4 presents this data from the Google mobility report. We note a strong decrease of all types of mobility before, and shortly after the lockdown. Afterwards, the mobility stays very stable, but an additional decrease can be seen coinciding with the second lockdown as all-non essential mobility was closed. This additional reduction of mobility during the closure of all non-essential is better illustrated in Fig. S5, which shows the average mobility during the three subsequent lockdowns. We also note that the third lockdown shows a slightly stronger mobility reduction compared to the first one despite the same restrictions being applied.

Comparing the mobility reduction with the decrease in *R*_*t*_, we find an elevated cross-correlation. Figure S6 presents the cross-correlation between the mobility types and *R*_*t*_ for the CCAA. A detailed distribution for the maximal cross-correlation is shown in Fig. S7. Additionally, with very little exceptions, the maximum cross-correlation is reached without any lag between the two time series. The exception here is the mobility related to grocery shops and pharmacies. This negative lag is due to the strong increase in visits of grocery stores and pharmacies due to fear of supply shortages as shown in Fig. S4.

### Detailed analysis of segments in the national *R*_*t*_

In the main text we separated *R*_*t*_ into three linear segments: the “free” spreading phase before restrictions, the initiation of the epidemic response, and the lockdown. Here, we present a more detailed analysis regarding the epidemic response and the lockdown. Therefore, we cut the time series of *R*_*t*_ at the turning point as the decrease is initiated in the three segment analysis. Subsequently, we analyze the different segments observed until the end of lockdown. We choose the number of segments according to the Bayesian information criterion (*40*) and corroborated with the Akaike information criterion (*41*) by iterating from 1 to 15 segments.

The identified segments are shown in Fig. S8 (bottom) together with the evolution of mobility according to the COVID-19 Community Mobility report from Google (*42*) (top). Interestingly, between the first and second breakpoint, we observe a substantial reduction of *R*_*t*_ before mobility starts to decrease. The second breakpoint then coincides with the initiation of mobility reduction. The third breakpoint matches the implementation of the first lockdown on March 15. At breakpoint 4, *R*_*t*_ starts a plateau together with the mobility level. The breakpoint 6 then corresponds to the shutdown of all non-essential activity, the second lockdown. This also coincides with a further reduction in overall mobility as shown in Fig. S5.

The lift of the second lockdown and the associated mobility increase, seem not to have caused any increase in *R*_*t*_. It is probable that the good weather conditions, as well as an increased individual awareness, raised by the elevated death toll and reflected in the reduced mobility (see Fig. S5), have contributed to a continuous decrease in *R*_*t*_. Finally, at breakpoint 9 the increase in *R*_*t*_ coincides with the lift of the lockdown and corresponding increase in mobility. Various identified breakpoints cannot be associated with any nation-wide introduced measures put in place by the national authorities or changes in mobility. They may stem from measures introduced in the different CCAA.

### Sensitivity Analysis

For adjusting the model to the daily fatalities we assumed a gamma distribution. The mean of the distribution was fixed to 11 days according to the information provided by the health authorities (*30*). Furthermore, we adjusted the shape factor to match the interquartile range (7 and 17 days) since this is, besides the mean, the only information we have at our disposal of the empirical distribution. Even though gamma distributions have been shown to describe particularly well the time between symptom onset and disease (*31*), we performed the same analysis making use of a log-normal distribution. We assumed the same priors as in the case of the gamma distribution.

Adjusting the log-normal distribution to the interquartile range, we found a logarithmic standard deviation of 0.54. The log-normal distribution is compared to the gamma distribution in Fig. S9. The parameters we find adjusting to the daily fatalities are: I0 = 261 (CI: 139 – 434), R1 = 4.5 (CI: 4.0 −5.0), R2 = 0.732 (CI: 0.724 −0.740), BP = 2020-03-11 (CI: 2020-03-09 – 2020-03-12). The estimations of R1 and R2 lead to a doubling time and half-life time of 1.94 (CI: 1.75 – 2.11) days and 12.1 (CI: 11.7−12.5) days, respectively (*12*). The results are not substantially altered with respect to a gamma distribution. The smaller *R*_*t*_ before the epidemic leads to a higher number of initially infected individuals. Furthermore, the shape of the log-normal distribution causes *R*_*t*_ to decrease a day earlier. The posteriors and their correlations are shown in Fig. S10.

Figure S11 shows the obtained fit between the model and the data. Figure S12 shows the inferred form of *R*_*t*_, while in Fig. S13 we present the evolution of under detection. The figures show very similar results as in the main text. Regarding the counterfactual scenarios, we find 5, 274 (CI: 4, 471 – 6, 005) fatalities and an attack rate of 1.4% (CI: 1.1− 1.5%) for a 7 day earlier epidemic response. Considering a 7 day later epidemic response, the model shows 112, 416 (CI: 102, 537 – 125, 562) fatalities and an attack rate of 28.8% (CI: 26.2 −32.%). The complete results are shown in Fig. S14.

**Figure S1:**
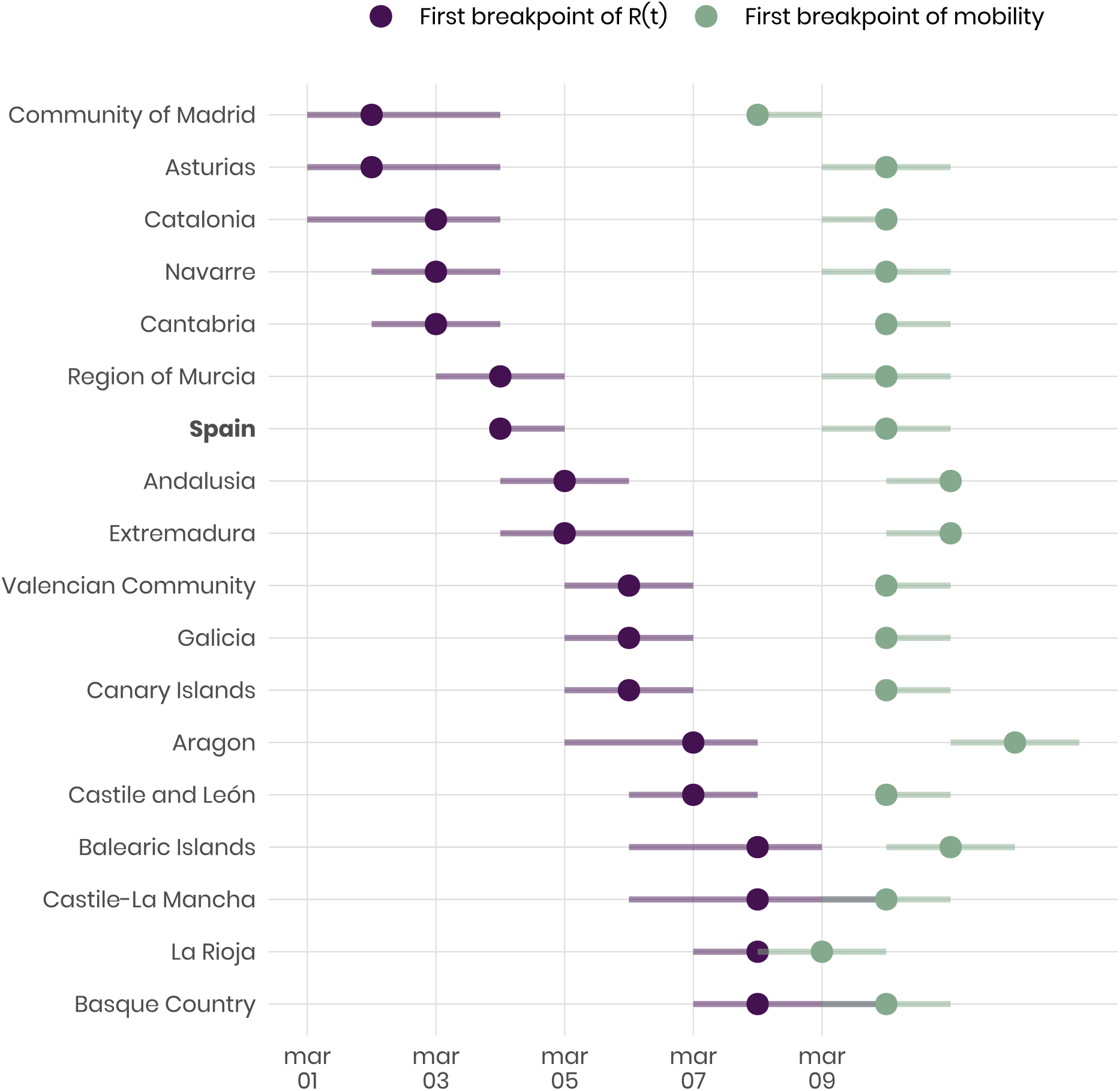
First breakpoint in *R*_*t*_ and workplace related mobility for Spain and all CCAA. Points indicate the mean, whereas horizontal bars indicate 95% credible intervals. We explicitly separated the time series into three segments, and identified them with the segmented package (*26*). Similarly as in the main text, we assumed the first segment of the time series was constant, and the others linear. To discard initial fluctuations in *R*_*t*_ due to a small incidence, we did not consider values before March 1. Additionally, we cut the series on March 28 to exclude the additional decrease in mobility due to lockdown 2. Overall, *R*_*t*_ decreases substantially earlier than mobility.

**Figure S2:**
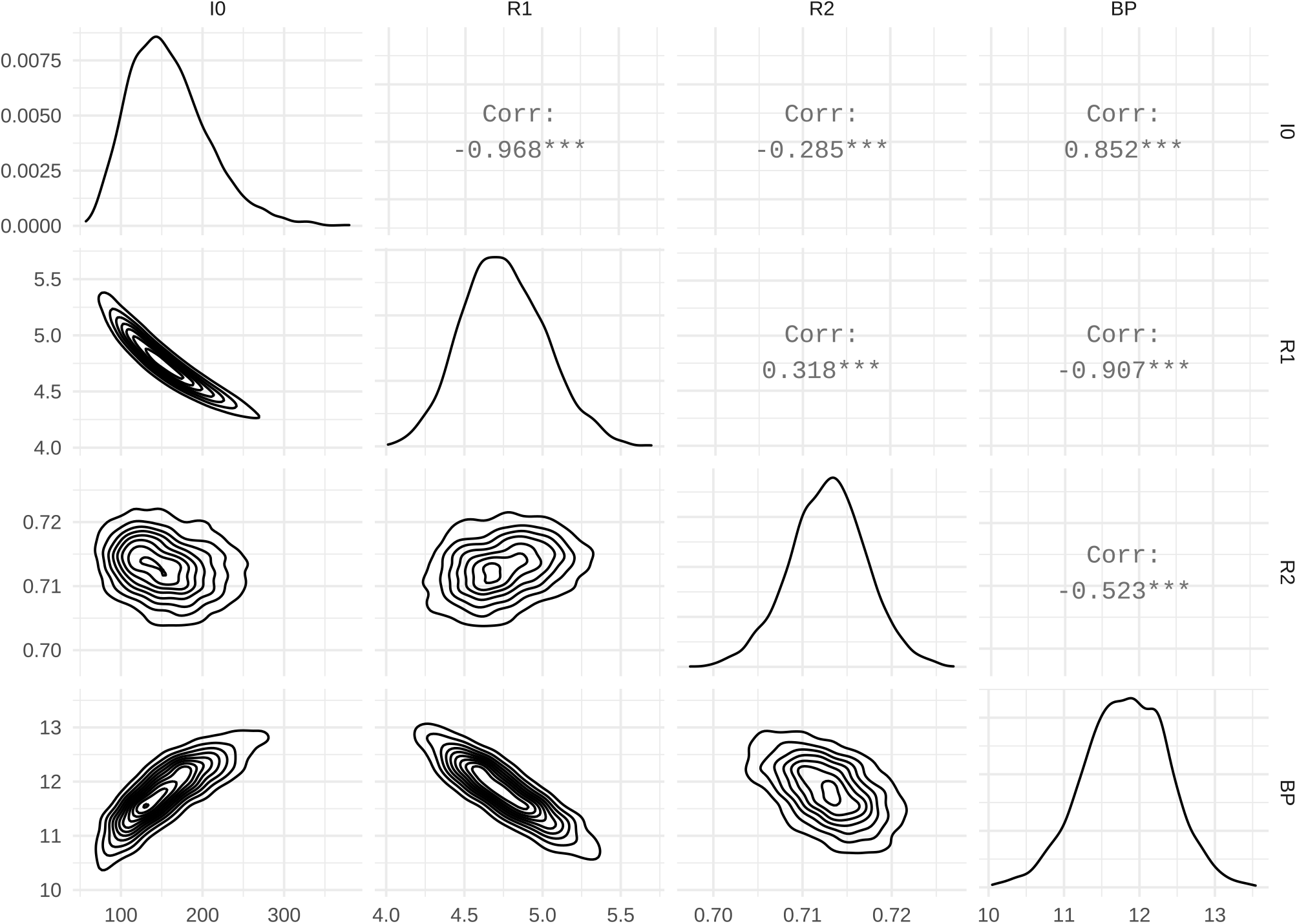
Posterior distribution of the model parameters and their correlations, assuming the time between symptom onset to death follows a gamma distribution. The range of the flat priors was larger than the posteriors in all cases. For the variable BP, the integer values refer to the dates in March 2020.

**Figure S3:**
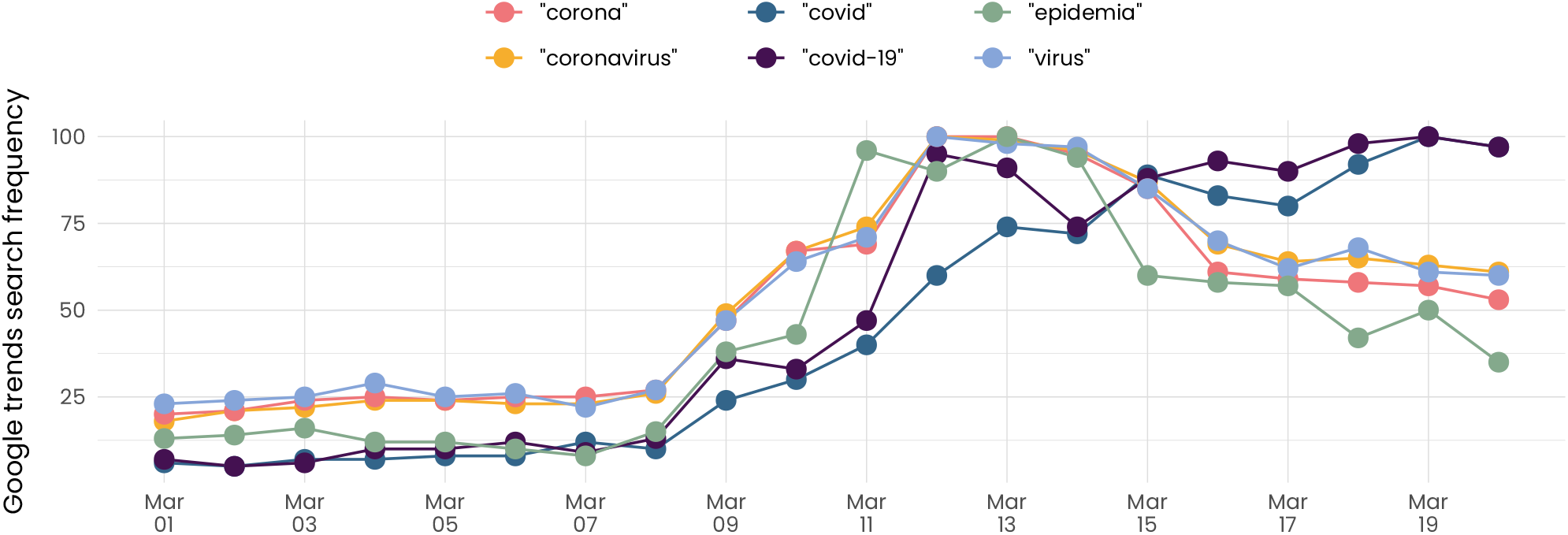
Frequency of different COVID-19 related search queries according to Google Trends. Note that Google Trends normalizes the frequency of queries between 0 and 100 during the considered period. All key words see a sharp increase from March 9 onward. On the same day, the Community of Madrid (*43*) closed all educational centres and the Basque Country (*37*) announced their closure for the next day. These closures correspond to the first NPIs introduced in Spain.

**Figure S4:**
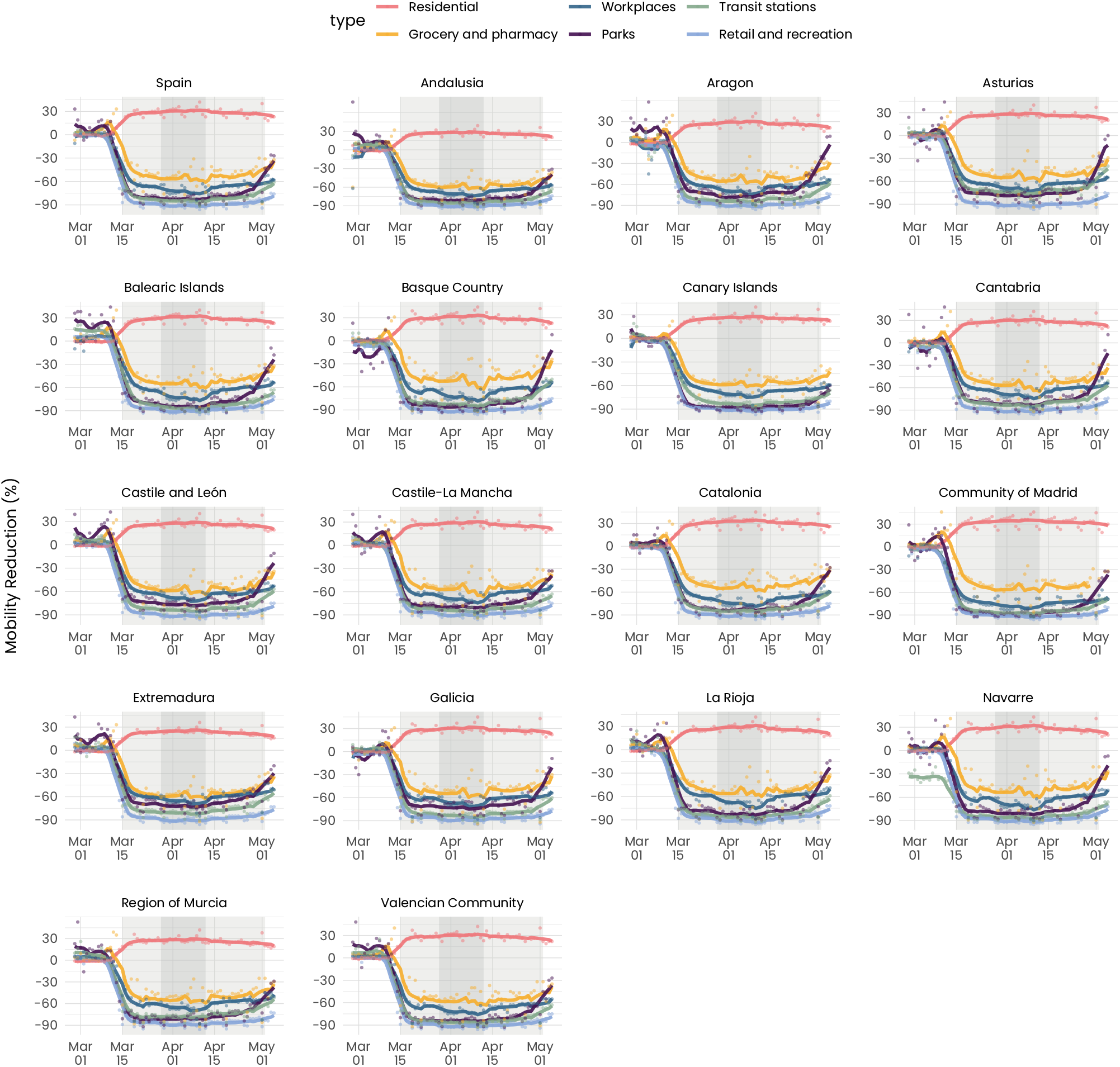
Visualization of the mobility data from the COVID-19 Community Mobility report from Google (*42*). Dots indicate the data points, while solid lines represent a centered, 7 day rolling average. The shaded areas in light gray indicate the lockdowns 1 and 3. In dark gray, we indicate the lockdown 2.

**Figure S5:**
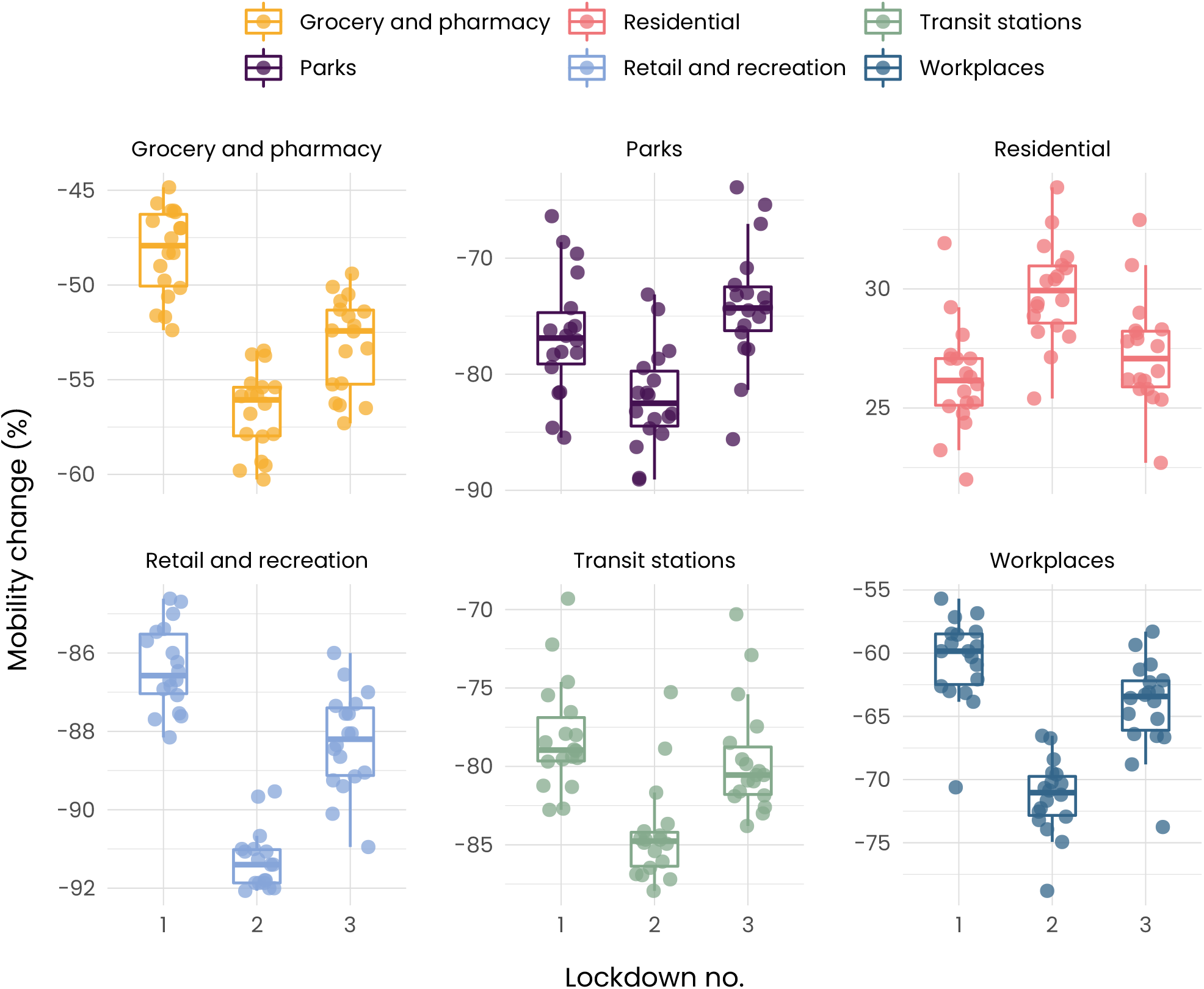
Average mobility according to the COVID-19 Community Mobility report from Google (*42*) during the three lockdown stages in Spain. Mobility is substantially lower during the second lockdown since all non-essential economic activity was shutdown. Additionally, mobility was lower during lockdown 3 when compared to lockdown 1, even though restrictions were the same. This indicates an increased awareness among the population, probably due to the huge death toll to this point.

**Figure S6:**
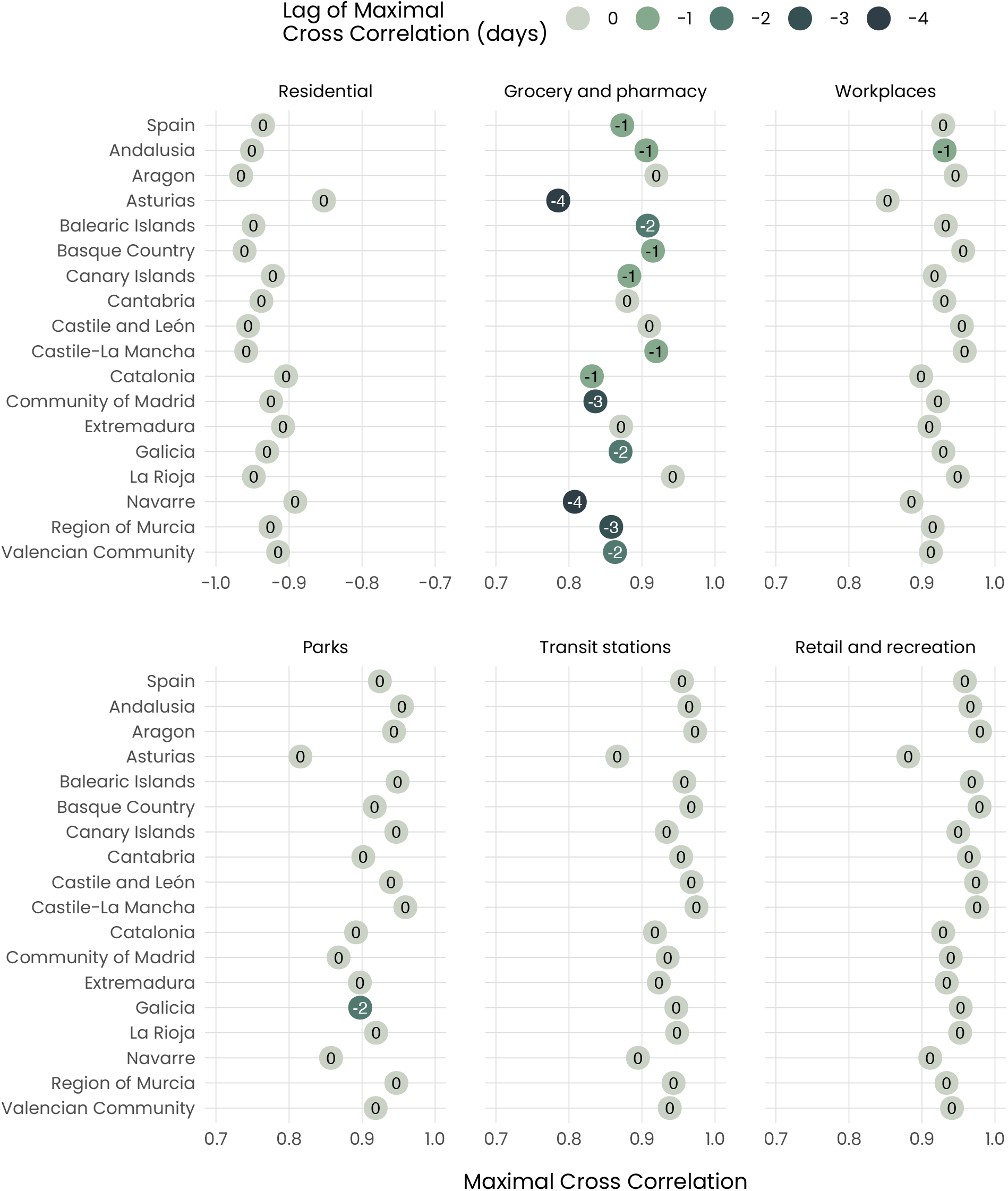
Lag and value of maximal cross-correlation between different mobility types, according to the COVID-19 Community Mobility report from Google (*42*) and *R*_*t*_. Negative lag indicates that *R*_*t*_ precedes mobility.

**Figure S7:**
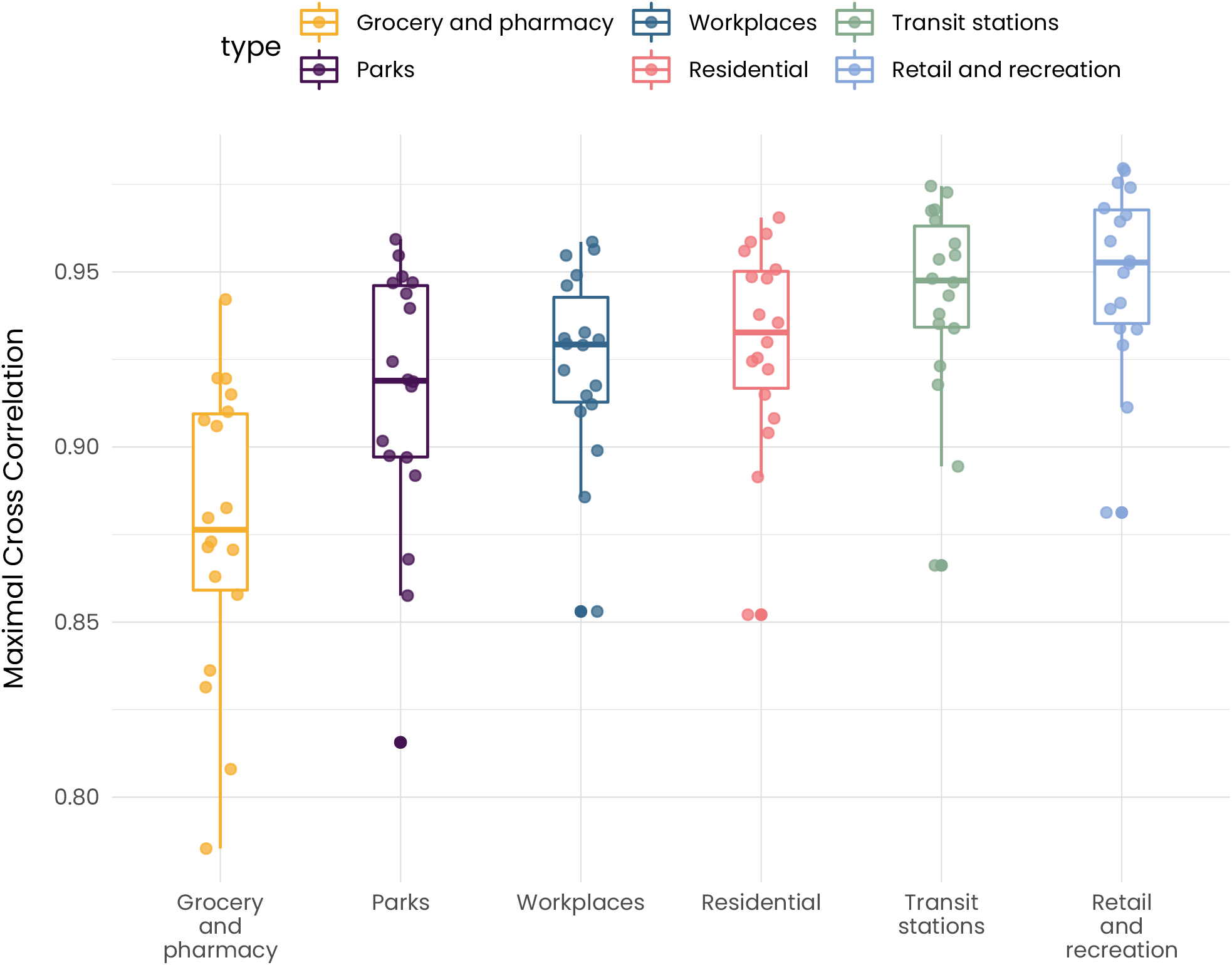
Maximal cross-correlation among CCAA between different mobility types, according to the COVID-19 Community Mobility report from Google (*42*) and *R*_*t*_. For the Residential mobility category we take the absolute value to facilitate visualization.

**Figure S8:**
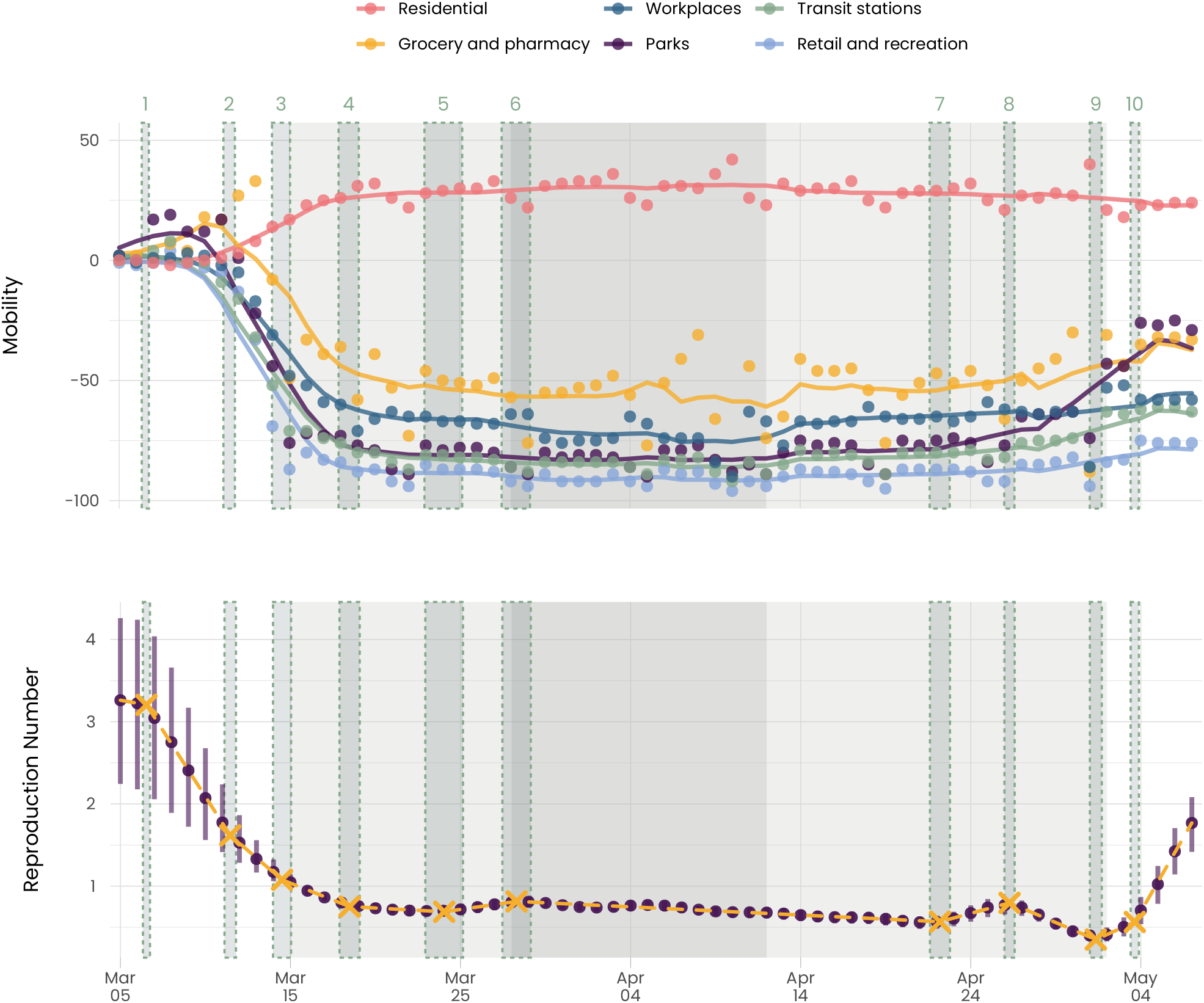
(Top) Average national mobility according to the COVID-19 Community Mobility report from Google (*42*). Dots indicate the data points and solid lines represent a centered, 7 day rolling average. (Bottom) *R*_*t*_ and the identified segments. Purple dots are the estimated values of *R*_*t*_ while bars indicate credible intervals. Yellow crosses refer to the breakpoints, and green dashed rectangles indicate their uncertainty range. The shaded area in light gray indicates lockdowns 1 and 3. In dark gray, we indicate lockdown 2.

**Figure S9:**
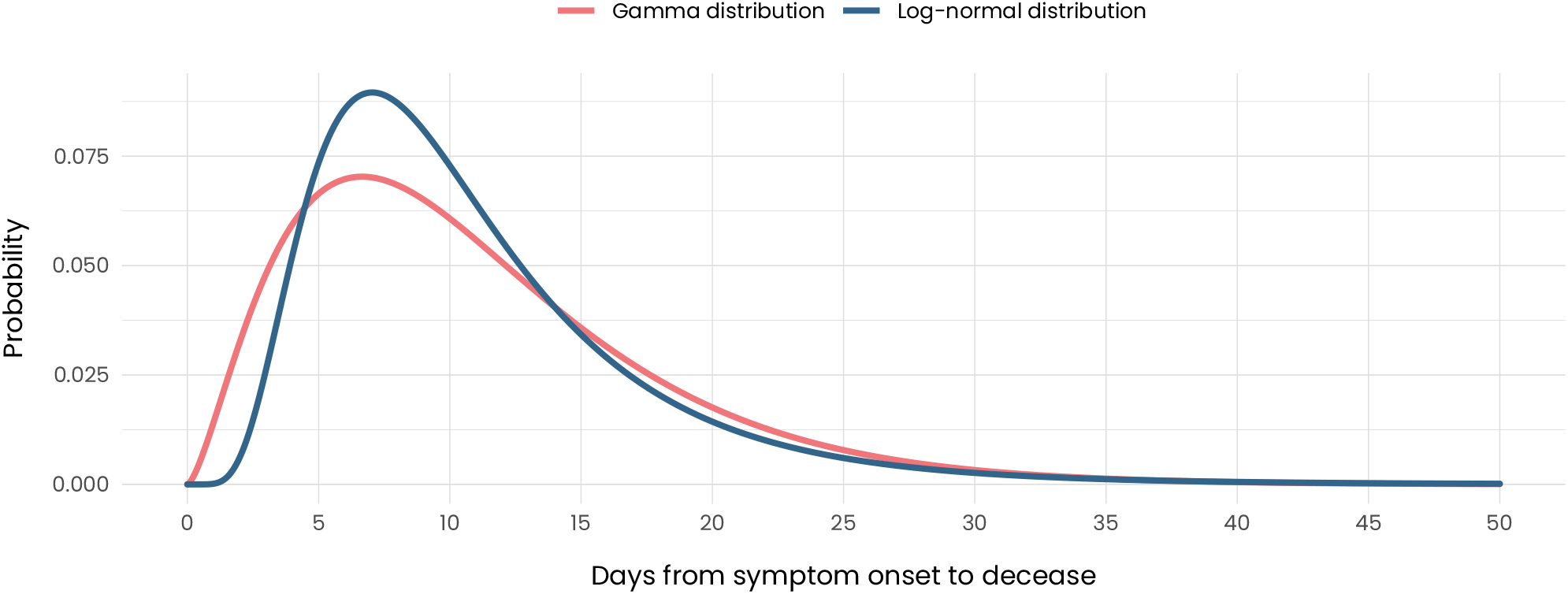
Gamma and log-normal distributions used for the time between symptom onset and decease. The average of both distributions is 11 days. The gamma distribution has a shape factor of 0.63, whereas the logarithmic standard deviation of the log-normal distribution is 0.54.

**Figure S10:**
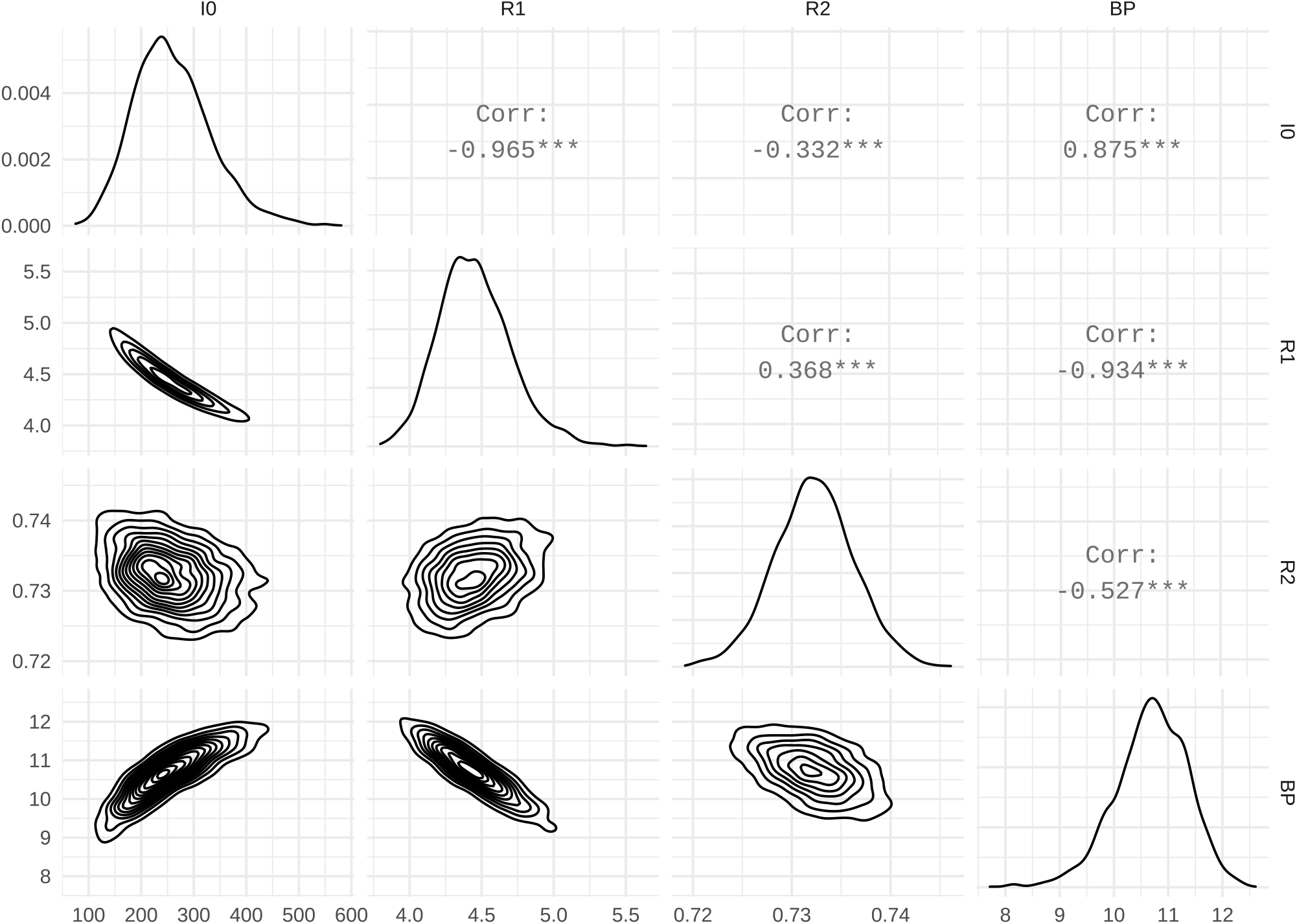
Posterior distribution for the model parameters and their correlations, assuming the time between symptom onset to death follows a log-normal distribution. The range of the flat priors was larger than the posteriors in all cases. For the variable BP, the integer values refer to the dates in March 2020.

**Figure S11:**
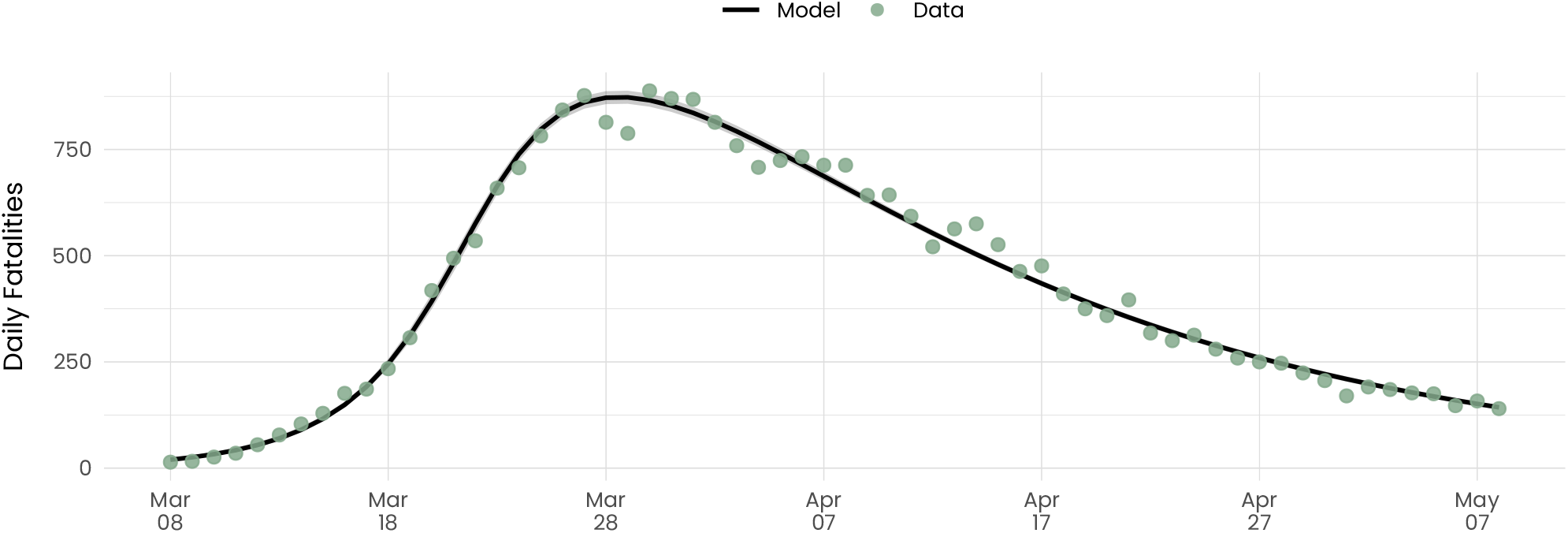
The daily fatalities in Spain from March 8 to May 8 compared to the daily fatalities for the sensitivity analysis. In the sensitivity analysis, we used a log-normal distribution describing the time between from symptom onset to decease. The shaded area represents 95% credible intervals.

**Figure S12:**
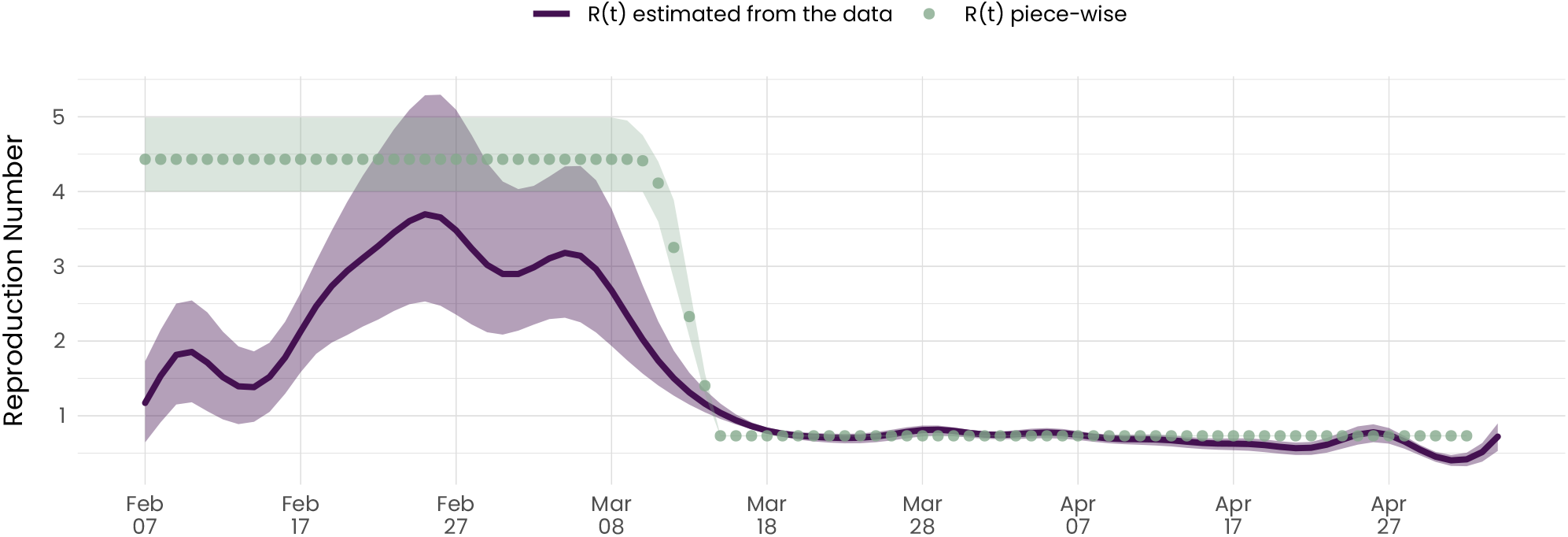
*R*_*t*_ estimated from the reconstructed exposure times vs. the model inferred as part of the sensitivity analysis. This is equivalent to Fig. 4A in the main text when using the log-normal distribution.

**Figure S13:**
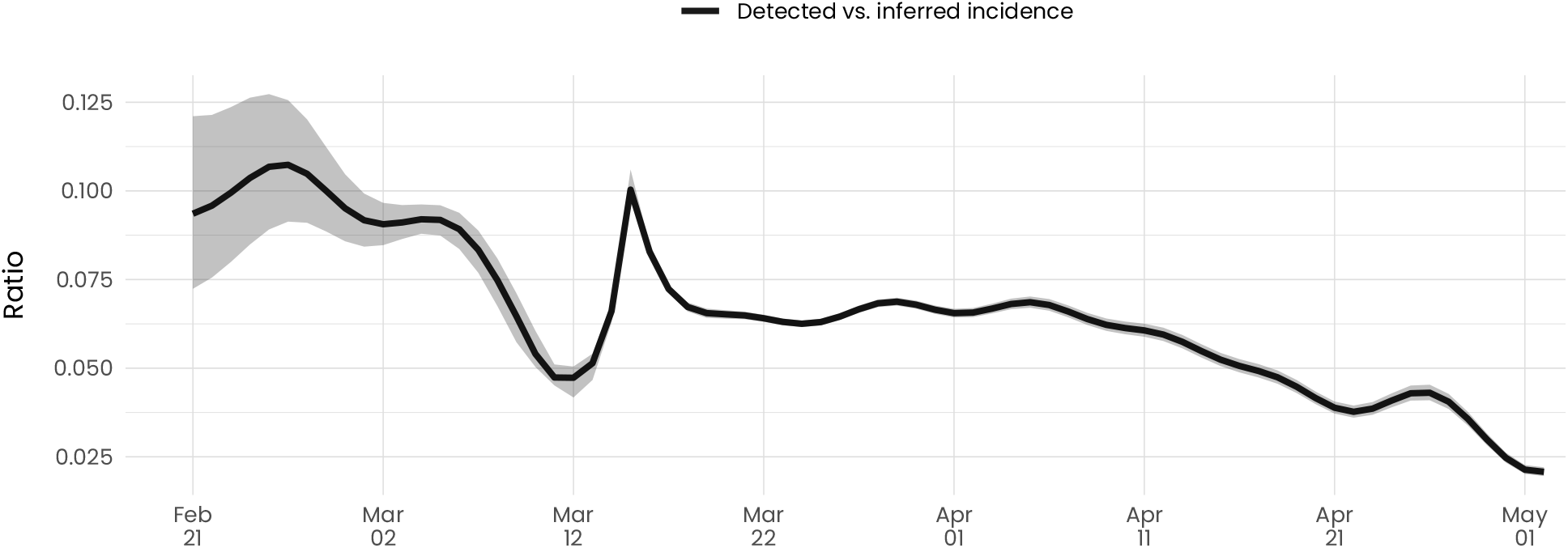
Ratio between the reconstructed exposure times and the model inferred infections as part of the sensitivity analysis. This is equivalent to Fig. 4B in the main text when using the log-normal distribution.

**Figure S14:**
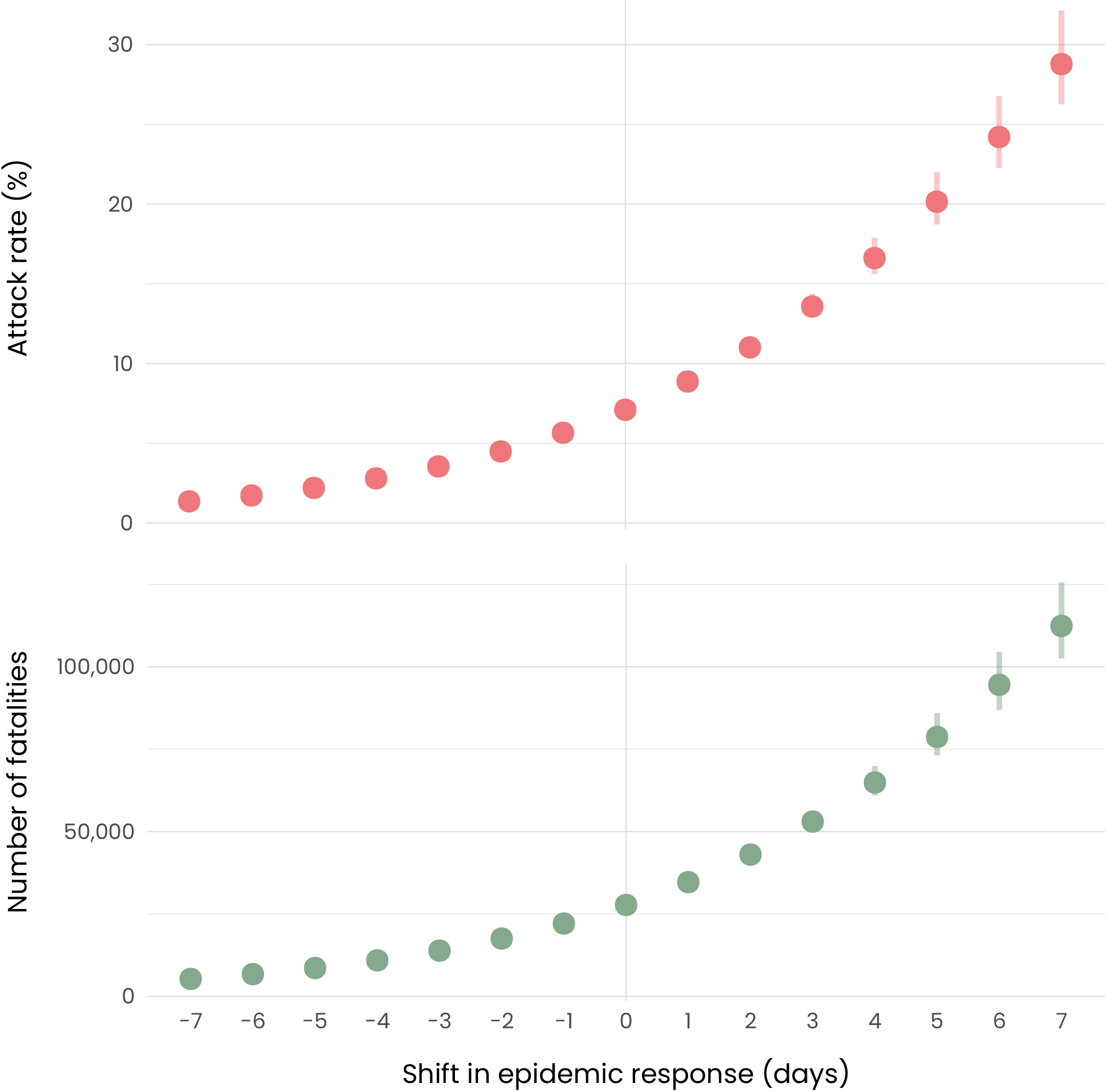
Counterfactual scenarios for the attack rate as well as the total number of fatalities. This is equivalent to Fig. 4C in the main text when using the log-normal distribution.

